# Structure-Function Correlation Using a Targeted Image-Guided Microperimetry Approach for Retinal Atrophic Diseases: A Methods Study

**DOI:** 10.1101/2025.07.07.25330852

**Authors:** Rubbia Afridi, M Sohail Halim, Mohamed Ahmed, Hikmet Yucel, Zoha Zahid Fazal, Kholood Janjua, Mauro Campigotto, Syed Mahmood Shah, Quan Dong Nguyen, Yasir Jamal Sepah

## Abstract

**Purpose:** This proof-of-concept aimed to evaluate the feasibility of targeted, image-guided microperimetry (IGMP) by integrating Optical Coherence Tomography (OCT) and Fundus Autofluorescence (FAF) for lesion mapping and functional assessment in retinal atrophic diseases.

**Methods:** 22 eyes were identified, of which 17 had early geographic atrophy (GA) while 5 had Stargardt Disease (SD). Disease transition zones (TZs) on OCT and FAF were annotated onto enface infrared images. These were then exported with microperimetry assessments to custom MATLAB applications. IGMP patterns were thereafter created and imported using MAIA and Nidek microperimeters for subsequent scans. Mean procedure and examination times (mins), retinal sensitivity (dB), approach feasibility, and algorithm compatibility of targeted IGMP were reported as outcomes.

**Results:** Targeted IGMP was quicker than the standard 10-2 grid for both SD and GA lesion assessments. SD cases showed longer mean procedure and examination times and lower mean retinal sensitivity in all zones compared to GA lesions. Targeted IGMP procedure was lengthier for Nidek (29.2±7.1 mins) than MAIA (17.4±4.3 mins) although examination took longer on MAIA (4.0±1.0 mins) than on Nidek (3.6±1.2 mins). Overall, IGMP approach is feasible (100.0%) and the algorithm is compatible (100.0%) on both MP devices for all subjects tested.

**Conclusions:** This study establishes the feasibility of an IGMP workflow in providing targeted functional assessments in retinal degenerative diseases. Our approach may help enhance structure-function correlation and optimize the efficiency of microperimetry integration for clinical trials. Further validation in larger cohorts is needed to assess its broader clinical applicability.

## Introduction

The development of effective ophthalmic therapies requires reliable biomarkers that accurately reflect disease progression and treatment response. For retinal degenerative diseases characterized by localized and progressive damage, a robust alignment between retinal structure and visual function is especially important^1^. Best-corrected visual acuity (BCVA)^2^ and lesion size^3^ have previously been used as endpoints for clinical trials investigating inherited retinal diseases (IRD) such as early geographic atrophy (GA) and Stargardt disease (SD). However, since BCVA often correlates weakly with disease progression^4,5^ and visual function^6–8^, the need for more sensitive functional tests was recognized by the National Eye Institute (NEI) in 2017^9^. **Figure 1** demonstrates how central visual performance may fail to reflect extensive structural damage in many macular diseases, especially in cases with no or complete foveal involvement. ^1,8^.

**Figure 1:**
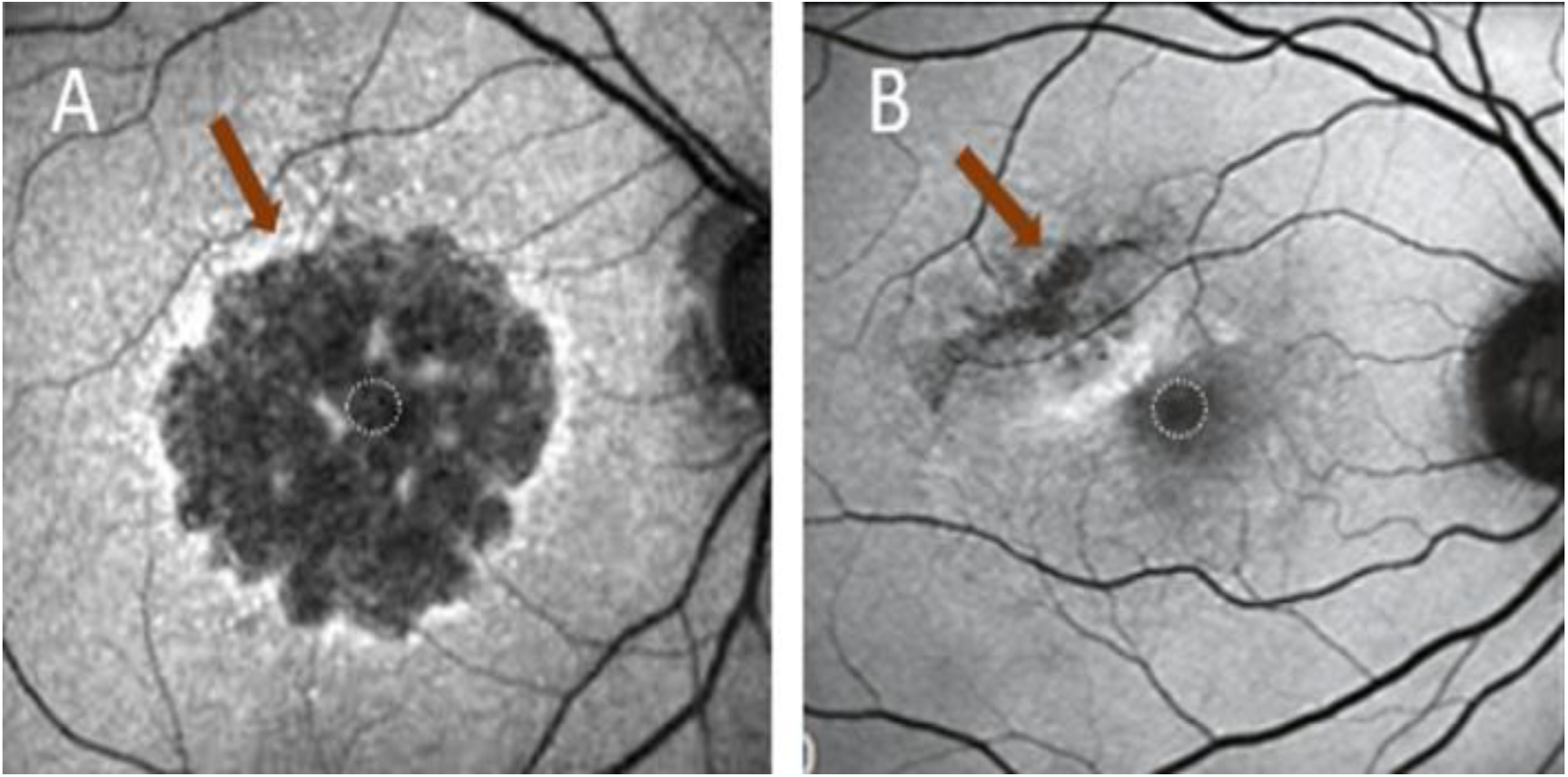
Irreversible damage of atrophic lesions located in foveal **(A)** or extrafoveal **(B)** region can influence the potential observable change in visual acuity with treatment.

Post-hoc analyses from recent clinical trials, such as OAKS & DERBY^3^ and Chroma & Spectri^10^ studies, have thus reported microperimetry (MP) assessment using the 10-2 protocol a more sensitive biomarker for retinal sensitivity decline and structure-function correlation in IRDs^11,12^ as illustrated in **Figure 2**. Additionally, **Figure 3** by Chang et. al^13^ shows that the most pronounced decline in retinal sensitivity often clusters along the transitional zone (TZ) in the macular region. These findings emphasize the importance of targeting TZ to capture early, spatially specific functional loss. However, the application of MP has significant limitations. The extensive time for each assessment, often exceeding 20 minutes per eye at baseline^14–16^, along with the induced patient fatigue^9,17^ could impact the validity of results. In addition, standard grids cannot be adapted to the size or location of lesions, frequently including regions of irreversibly damaged retina and thus limiting specificity. These constraints underline the need for a refined approach that enhances efficiency, reduces patient burden, and improves correlation with structural biomarkers.

**Figure 2:**
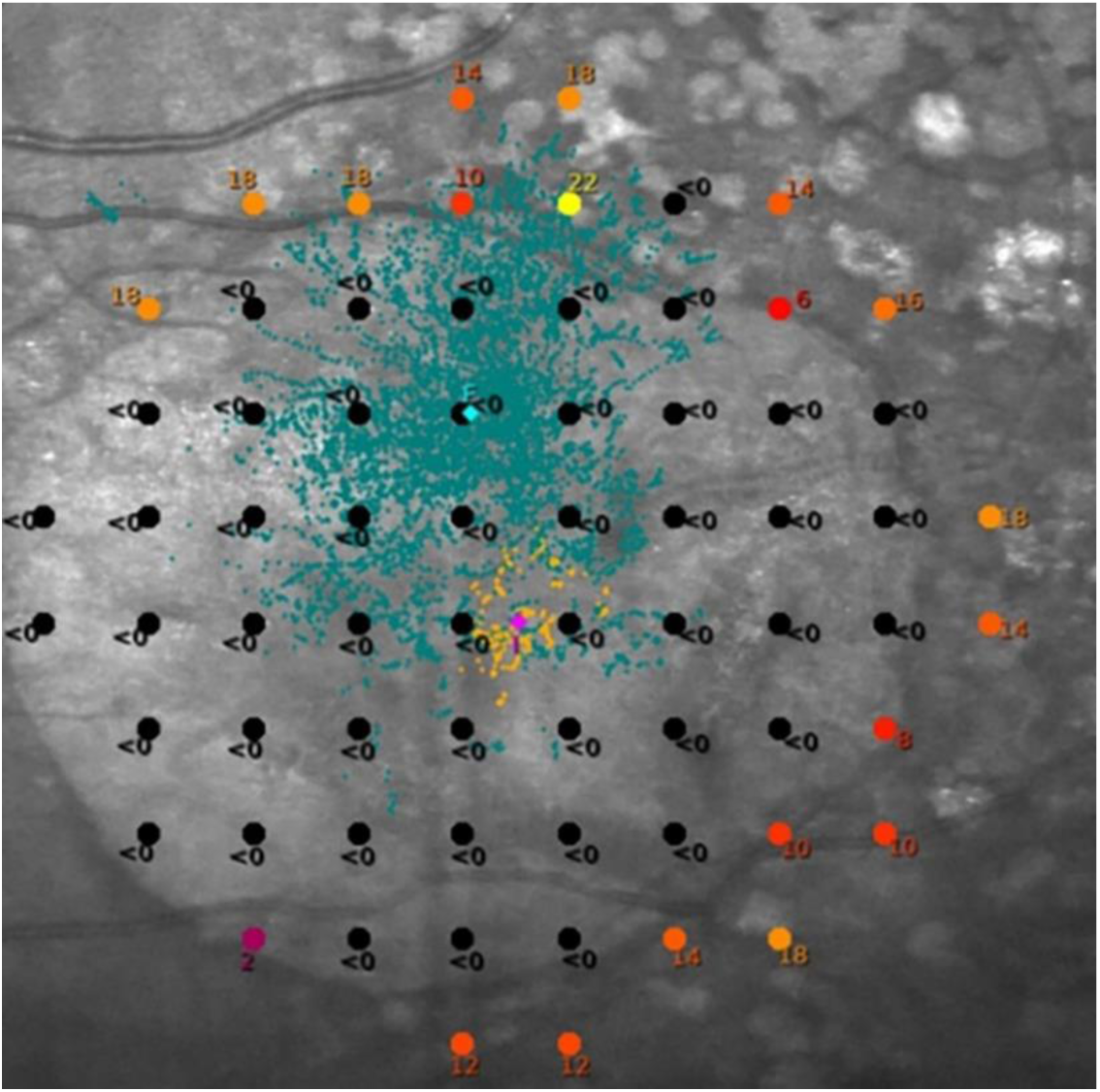
Representative microperimetry scan using a 10-2 scan pattern demonstrating a large area of scotoma.

**Figure 3:**
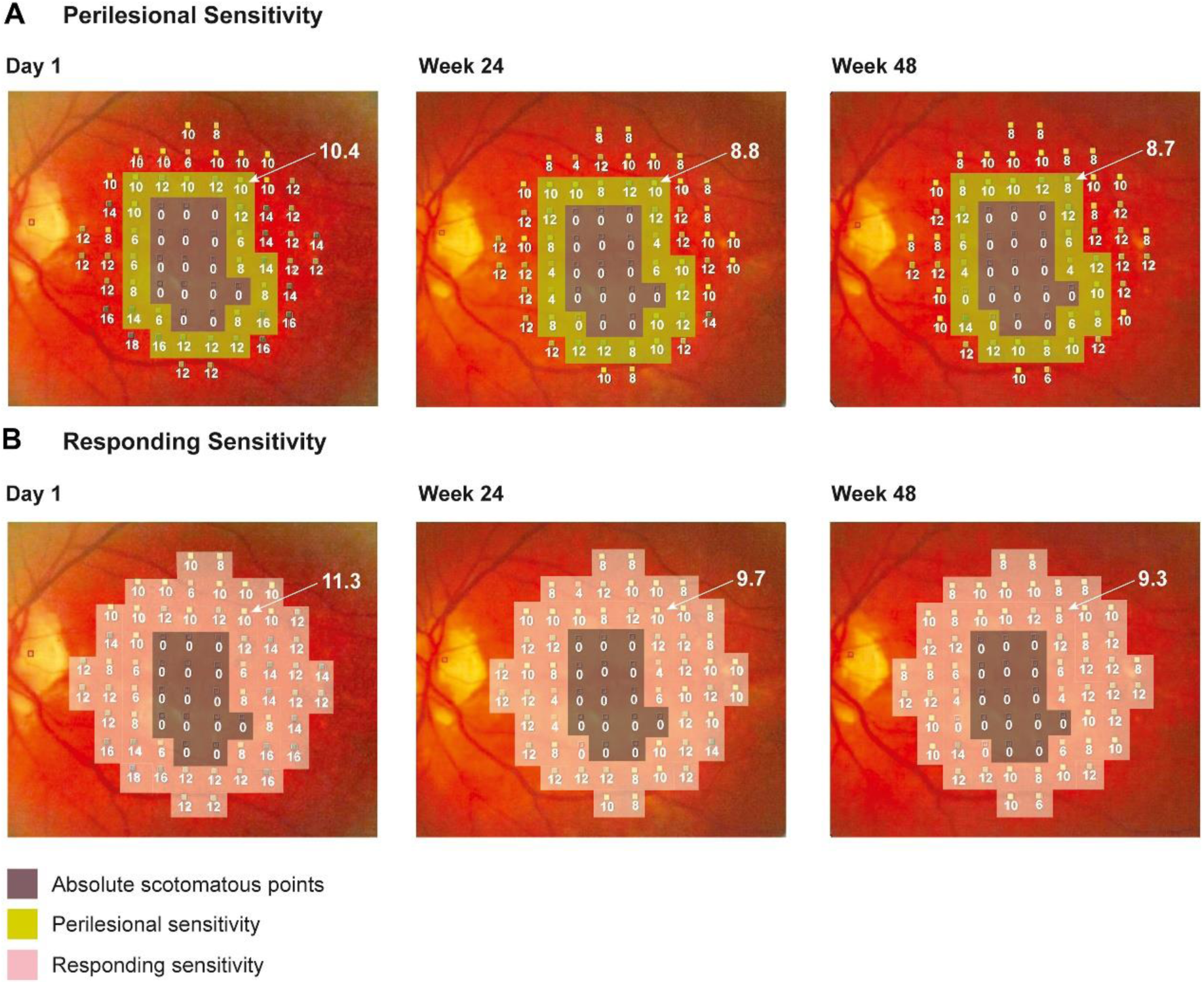
Image from Chang et. al^13^ reprinted with permission demonstrating absolute scotoma points at the baseline visit and the subsequent identification of two regions of interest, perilesional area **(A)** and responding area **(B)** outside of the absolute scotomas.

In response to these challenges, our team has developed a novel, image-guided microperimetry (IGMP) method that integrates gold standards of structural imaging, i.e. fundus autofluorescence (FAF) and optical coherence tomography (OCT), to delineate atrophic lesions from TZ and healthy retina with high precision. This approach allowed us to adapt test grids to anatomically relevant regions, enhancing the accuracy of functional assessment. Our goal was to improve the efficiency of testing while ensuring a robust correlation between structural and functional data using the previously defined and innovative speeded up robust features (SURF) approach^18^. In terms of its translational relevance, the custom IGMP algorithm could help improve MP integration into clinical trials for better understanding of disease prognosis and allow its use as a biomarker for disease progression and response to therapeutic agents for retinal degenerative diseases.

## Methods

This section details the imaging and microperimetry protocols utilized on patients with IRDs, specifically GA and SD. Each step has been outlined to facilitate clarity and encourage precise execution of the protocol. With adherence to the clearly defined steps detailed in this methodology, researchers can replicate the study and explore the functional changes in the retina with enhanced precision. **Figure 4** summarizes the workflow for this proof-of-concept and can be segmented into the **pre-microperimetry** and **microperimetry** phases.

**Figure 4:**
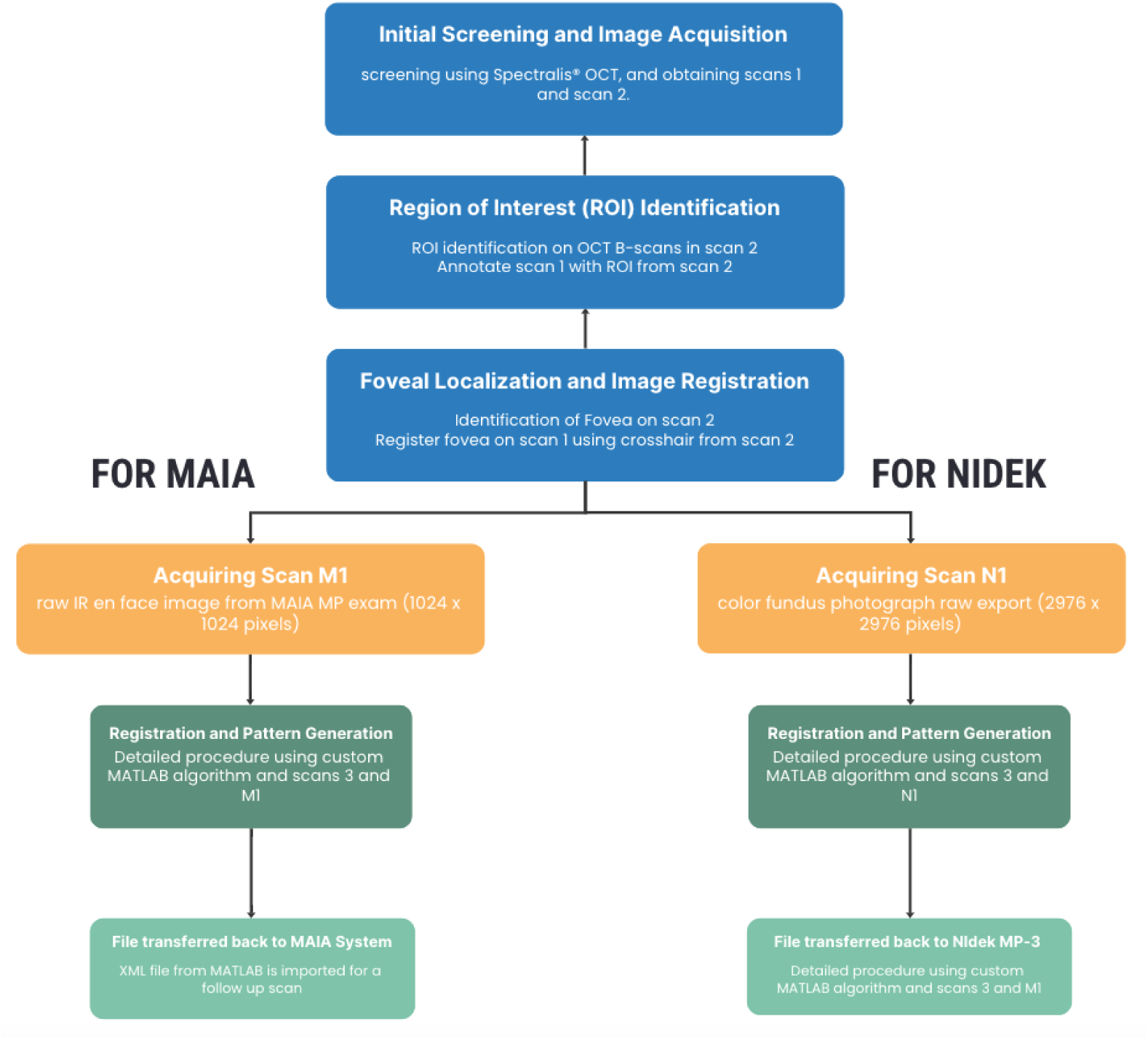
Summary of workflow for image-guided microperimetry using MAIA and Nidek-MP3/S for patients with inherited retinal disease.

### Facilities and Resources

To replicate the methodology, the following requirements must be met:

- Access to the specified OCT, FAF, and MP equipment for multimodal imaging.
- A certified retinal photographer or equivalent trained personnel for image acquisition.
- Proficiency in Matrix Laboratory (MATLAB) for the execution of the custom algorithm.
- Understanding of the SURF feature extraction process for accurate registration.
- Familiarity with an extensible markup language (XML) file generation and importation for MAIA and Nidek-MP3/S systems.

### Imaging Equipment

For this study, OCT and FAF were obtained using Heidelberg Spectralis (HS®, Heidelberg Engineering GmbH, Heidelberg, Germany). MP was acquired with Macular Integrity Assessment system (MAIA iCare, North Carolina, USA) and MP3/S (Nidek, Gamagori, Japan) microperimeters.

### PRE-MICROPERIMETRY PHASE PROTOCOL

The pre-microperimetry phase involves two steps:

1. OCT/FAF imaging
2. OCT/FAF segmentation

### OCT/FAF Imaging

*a. Initial Screening and Image Acquisition:*

- Participants undergo initial screening using the HS® OCT. Two scans are obtained:

- **Scan 1 (S1):** A high-resolution Infrared (IR) enface OCT/FAF image centered on the fovea (768x768 pixels).
- **Scan 2 (S2):** A Detailed Report containing the enface OCT image and the B-scan at the fovea (2640x2040 pixels).

**b. Region of Interest (ROI) Identification:**

- ROI is identified particularly on the OCT B-scans in S2.
- Annotations are made on the IR enface image of S1 based on the ROI from S2.

**c. Foveal Localization and Image Registration:**

- The fovea is identified on S2 using the cross-hair on the enface image.
- Annotations from S1 and S2 are registered to identify the foveal center on S1.
- The resulting image, **Scan 3 (S3)**, is an IR OCT/FAF image with burnt-in annotations (768x768 pixels).

### OCT/FAF Segmentation

• **For GA lesions:** A high-resolution OCT scan of the macular region was acquired. An expert image grader identified the boundaries of complete Retinal Pigment Epithelium (RPE) Outer Retinal Atrophy **(cRORA)** and transition zone **(TZ)** using B- scans. Two enface images with annotated boundaries were extracted, with the TZ defined as the region resulting from the subtraction of the inner boundary from outer boundary of TZ identified in each image. TZ outer boundary was identified as the junction between the healthy retina (all retinal layers intact) and the area of disorganized outer retinal layers and RPE. The inner boundary would delineate the junction between TZ and cRORA. **Figure 5** thus illustrates a progressive GA lesion segmented using OCT, each with enface images **(5A,5C)** and corresponding B-scans **(5B,5D)** to delineate atrophic region from TZ.

**Figure 5:**
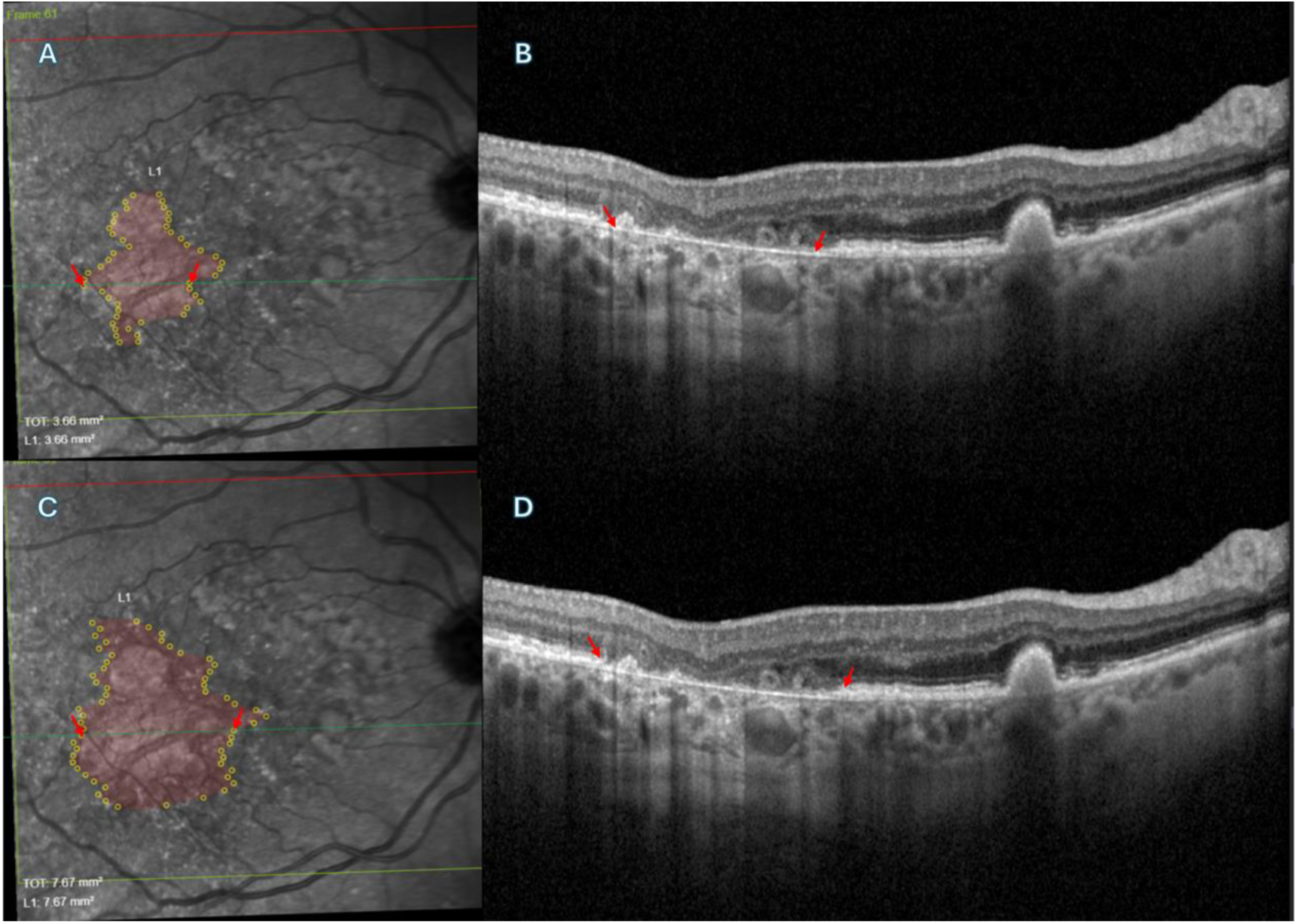
Example of OCT image segmentation as part of the pre-microperimetry protocol. **(A)** is an infrared image delineating enface representation of the GA lesion area with red arrows. **(B)** corresponds to lesion areas with complete retinal pigment epithelium (RPE) and outer retinal atrophy (cRORA) marked by red arrows. **(C)** illustrates the enface representation of the boundary between the morphologically non-atrophic retina and the transition zone. **(D)** shows a corresponding B-scan highlighting these boundaries with red arrows.

• **For SD lesions:** A comparable segmentation approach was employed for SD lesions as GA lesions; however, FAF imaging and segmentation was utilized instead to demarcate the atrophic lesion and TZ boundaries. The atrophic lesion was identified as the area exhibiting definitely decreased autofluorescence **(DDAF)**, equivalent to approximately 90% of the optic disc’s darkness level, aligning with the ProgStar^19^ definition. TZ was characterized as the area within 500 microns of the DDAF margin with autofluorescence reduction intermediate to that of healthy retina and DDAF, equivalent to approximately 50- 90% of the optic disc intensity, and consistent with the questionably decreased autofluorescence **(QDAF)** definition established by the ProgStar studies^19^. Despite its normal appearance, TZ—particularly on the temporal side—shows markedly reduced sensitivity compared to the healthy retina and may indicate a direction of future atrophic progression. The preserved and homogenous autofluorescence outside this TZ corresponds to healthy retina. Retinal sensitivity in these outer areas is generally higher than within DDAF and QDAF but still demonstrates significant functional loss.

### MICROPERIMETRY PHASE PROTOCOL

The microperimetry phase involves two steps:

1. Image acquisition
2. Registration and pattern recognition

Both steps can be executed using two different systems:

a. MAIA
b. Nidek-MP3/S

### Image Acquisition by MP

• **Standardized Pattern Acquisition:** A simplified 10-2 MP grid centered on the preferred retinal locus (PRL) was utilized to capture microperimetry exams using both MAIA and MP3. The raw data was exported from both devices.

• **Targeted Pattern Acquisition:** To generate targeted patterns, custom MATLAB applications for MAIA and Nidek-MP3/S were developed, which incorporated information from OCT and MP. The IR image from MAIA raw data or colored image from Nidek-MP3/S was registered to the annotated enface image from HS® using an interactive registration process. Fovea was identified using the extracted fovea report from HS®. The user would then proceed with the creation of the targeted MP pattern by placing markings around the lesion, along the eight radial axes in the TZ and healthy retina. The pattern was then saved. The MATLAB application can save a .tgz file for MAIA and the raw data format of Nidek- MP3/S. The modified .tgz file for MAIA was imported back in the device using the ‘append’ function. The targeted MP pattern was then displayed on the device and a follow-up could be performed to assess the retinal sensitivities in the ROI. A similar process was used to import the raw data for the Nidek-MP3/S device 6B. All scans were performed using Goldmann III stimulus size using a 4-2 strategy. **Figure 6** depicts how targeted IGMP on OCT delineates atrophic area from TZ and healthy retina for GA lesions by manual placement of MP test points with four points marking ring 1 (R1), eight points marking ring 2 (R2), and eight points marking ring 3 (R3), respectively. While **Figure 6A** illustrates how R1 demarcates the atrophic lesion, **Figure 6B** demonstrates the computed region of atrophic lesion (R1) and transition zone (R2) with R3 bordering TZ or R2 on the inner boundary and healthy retina on the outer boundary. Similarly, **Figure 7** represents how targeted IGMP on FAF can demarcate DDAF and QDAF for SD lesions by manual placement of MP test points with four points marking R1, eight points marking R2, and eight points marking R3, respectively. While the red boundaries in both images delineate DDAF, the region outside DDAF and within the green boundary, as shown in **Figure 7A**, or the yellow boundary, as shown in **Figure 7B**, representing QDAF or TZ synonymously for each imaged lesion.

**Figure 6:**
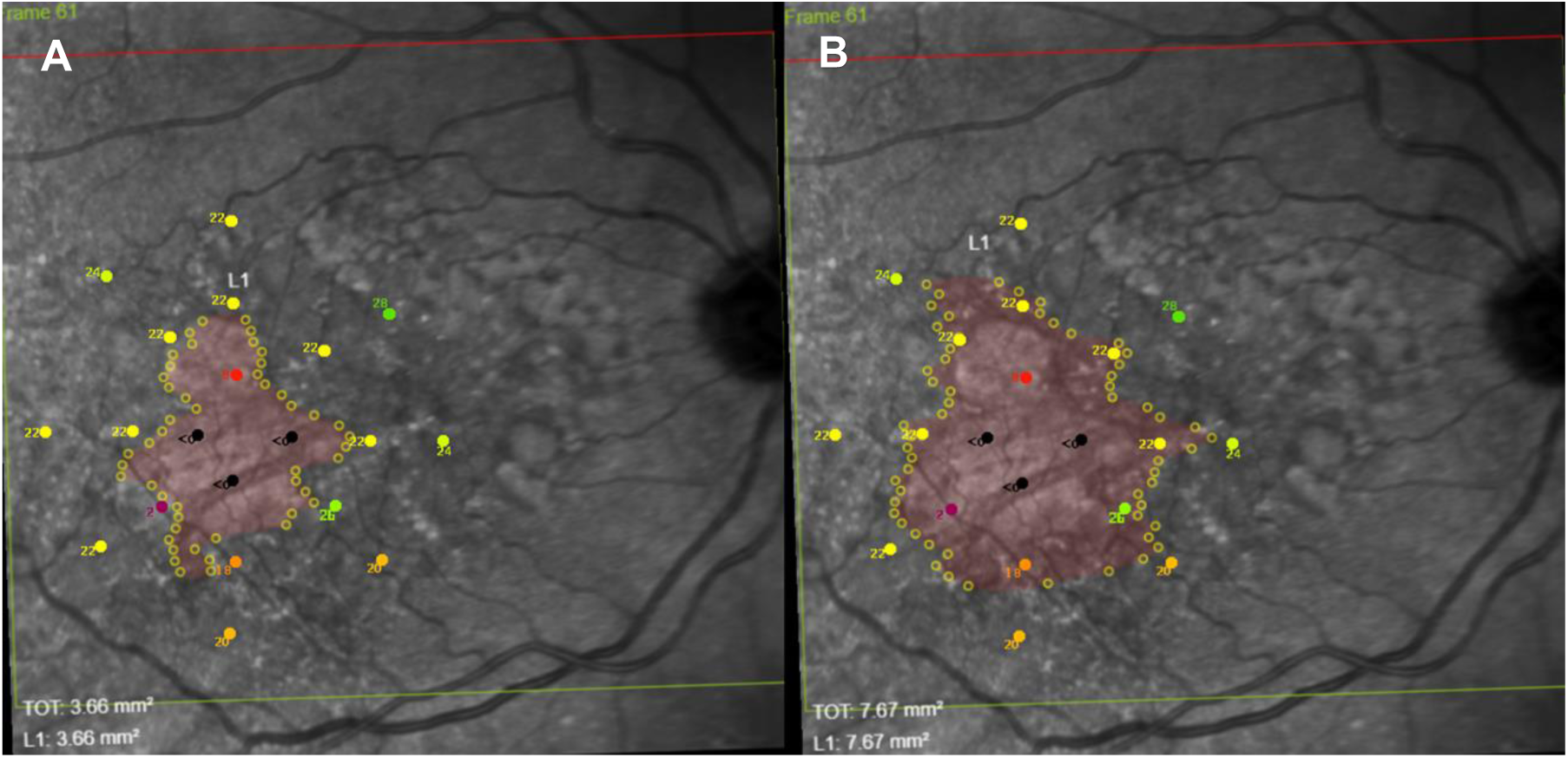
A representative targeted image-guided microperimetry pattern, illustrating the delineation of geographic atrophy (GA) boundaries on an infrared (IR) image **(A)** and the identified combined area of GA and transition zone **(B)**. MP test points were manually placed, with four points placed within the GA region (innermost points), eight within the transition zone (middle points), and eight in the unaffected retinal tissue (outermost points).

**Figure 7:**
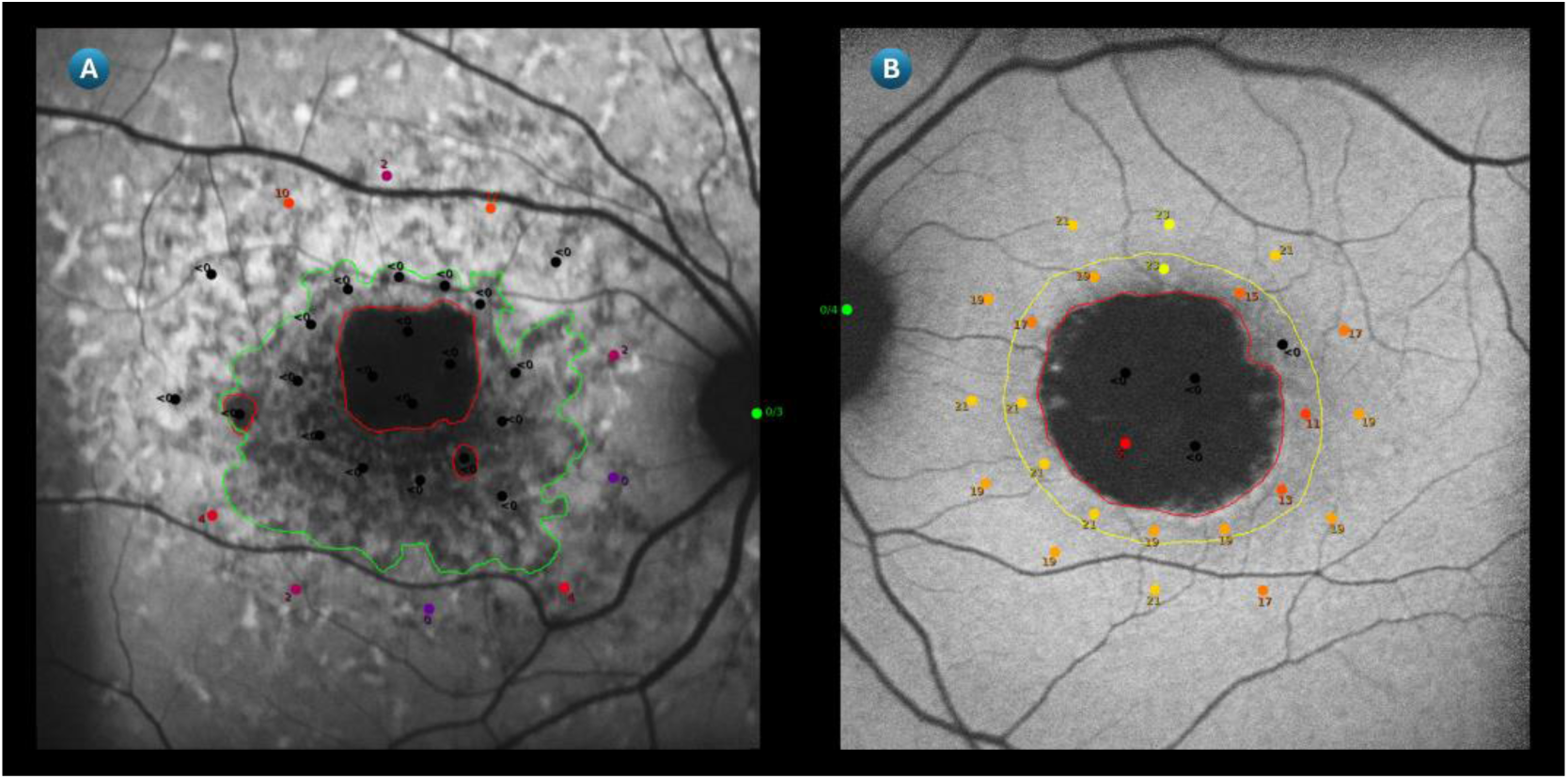
Representative images of targeted microperimetry in two eyes with Stargardt disease. In both panels, areas of Definitely Decreased Autofluorescence (DDAF) are outlined in red. **(A)** The region outside the DDAF and within the green boundary represents an area of Questionably Decreased Autofluorescence (QDAF). **(B)** The region outside the DDAF and within the yellow boundary corresponds to a transitional zone or QDAF.

**Figure 8:**
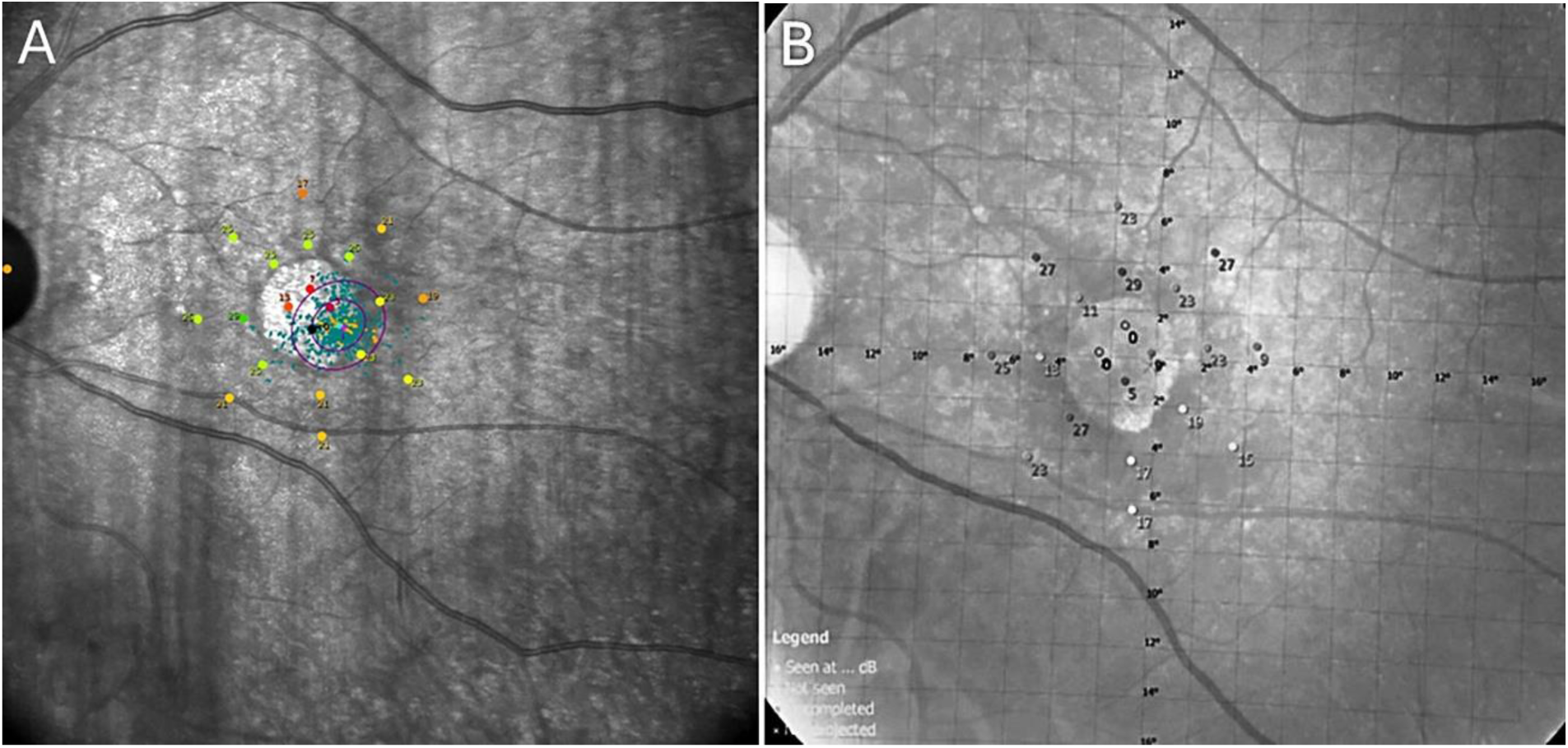
A representative targeted microperimetry scan of subject with geographic atrophy using MAIA **(A)** and Nidek- MP3/S **(B)**

### MAIA Microperimetry

**1. Image Acquisition:**

• A raw IR enface image is acquired after conducting the MAIA MP exam (1024x1024 pixels), referred to as Scan M1 (SM1).

**2. Registration and Pattern Generation:**

- Custom MATLAB algorithm is developed for image registration and XML pattern generation.
- The algorithm uses S3 and SM1 for processing.
- SM1 is registered to S3 using a GUI.
- The algorithm extracts and matches SURF features to compute a non-rigid transformation with a matching threshold of 55.
- The transformation is applied to S3, and the GUI is used to identify MP testing points on the transformed S3.
- Additional points are placed on vascular landmarks for accurate and precise grid alignment.
- The points’ coordinates are converted to polar coordinates and transcribed into an XML file which can be read by MAIA.
- The XML is imported into MAIA to align the testing grid, and a new custom exam is conducted.

### Nidek-MP3/S Microperimetry

**1. Image Acquisition:**

- Screening microperimetry is performed using Nidek-MP3/S with a 4-2 staircase strategy and Goldmann III stimulus size.
- A CFP is obtained from Nidek-MP3/S as raw export (2976x2976 pixels), referred to as Scan N1 (SN1).

**2. Registration and Pattern Generation:**

- The MATLAB algorithm requires S3 and SN1.
- Manual registration identifies landmarks on S3 and SN1 for alignment.
- Identified points for MP testing are marked on the registered S3 using GUI.
- Cartesian coordinates of these points are converted to polar coordinates using MATLAB.
- The Nidek-MP3/S calculates Perimetric Coordinate System (PCS) and Medium Coordinate System (MCS) coordinates, saving them as XML files.
- The new "mp_d_oi.xml" file contains the transformed coordinates and replaces the existing file in the Nidek-MP3/S raw data.

The modified data is imported back into the system for a follow-up MP session.

### Ethical Considerations

This study was approved by the institutional review board of Stanford University (IRB no. 68008) and adhered to the Declaration of Helsinki. Informed consent was obtained from enrolled subjects, and their protected health information was de-identified before storage in password-protected browsers accessible to the immediate study team only.

### Study Eligibility

Subjects fulfilling the following criteria were enrolled in the study:

- **Inclusion Criteria:** Age above 18 years for SD patients, age above 50 years for GA patients, previous diagnosis of an IRD (GA/SD), patient informed consent to study participation, sufficiently clear ocular media, adequate pupillary dilation and fixation.
- **Exclusion Criteria:** History of other ocular diagnosis, trials, or procedures in the last 3 months for either eye/ BCVA below 20 over 200/ high myopia > 6 diopters/ poor quality imaging/ risk of harm or injury to the patient.

### Study Procedures

17 eyes with GA and 5 eyes with SD were identified for this study. Of these, 11 eyes with GA and all 5 eyes with SD were initially screened using MAIA. Owing to the delay in the development of customized solution for targeted IGMP on Nidek-MP3/S, zero eyes with SD and only eight eyes with GA could be tested on Nidek, of which two were from the MAIA cohort. The same Nidek cohort was thereafter used for testing and comparing custom IGMP pattern feasibility and inter-operator compatibility on both MP devices. At screening, a complete history, physical examination, and vital signs were recorded for each patient after attaining informed consent. Adequate fixation and dilation were ensured for each eye examined as part of this study.

### Study Outcomes

For subjects examined as part of this methodology, the following variables were recorded:

- **Demographics:** retinal disease subtype (GA/SD), subject identification number, age in years, sex (Male/Female), race (White/Black/Unknown), ethnicity (Non-Hispanic-Non-Latino/Hispanic-Latino/Unknown), eye laterality (OD/OS).
- **Outcome Variables:** procedure time in minutes (mins) constituting of screening image acquisition time (mins) and custom pattern generation time (mins), examination time (mins) for targeted IGMP pattern versus standard 10-2 grid, and retinal sensitivity in decibels (dB) taken as an average of all points in each ring (R1, R2, R3) for custom pattern. Additionally, comparisons were drawn between Nidek and MAIA for total procedure time (mins), total exam time (mins), feasibility of approach (yes/no), and compatibility of algorithm (yes/no) for custom IGMP assessment.

### Statistical Analysis

Descriptive statistics were calculated using Microsoft Excel and reported as part of this study.

- **Categorical Variables:** Frequency and percentage were calculated.
- **Nominal Variables:** Mean and standard deviation (σ) were calculated.

## Results

This study included a total of 15 subjects, of which 12 (80%) presented with early GA and 3 (20%) had SD, respectively. All 3 SD subjects and 10 GA subjects were tested on the MAIA microperimeter, compounding to 13 patients (86.7%) in the MAIA cohort. Only 4 subjects (26.7%) with GA could be tested on the Nidek microperimeter, of which 2 (GA-09 and GA-10) were previously tested on the MAIA device.

**Table 1** summarizes the microperimetry procedure time in mins, exam time in mins, and retinal sensitivity in dB for each lesion assessed in patients with SD or GA solely on the MAIA device as part of this study. For eyes with SD, the mean time for image acquisition was 15.1±1.9 mins while custom pattern generation took an average of 7.2±1.2 mins, accounting for a total mean procedure time of 22.3±3.1 mins. However, for eyes with GA, image acquisition took 5.8±1.5 mins while pattern generation took 10.4±1.5 mins on average. This implies that obtaining imaging was comparatively more time-consuming in SD patients while pattern generation took longer in GA patients. Overall, the mean total procedure time was shorter in GA patients, i.e. 16.2±2.1 mins, than in SD patients.

**Table 1:**
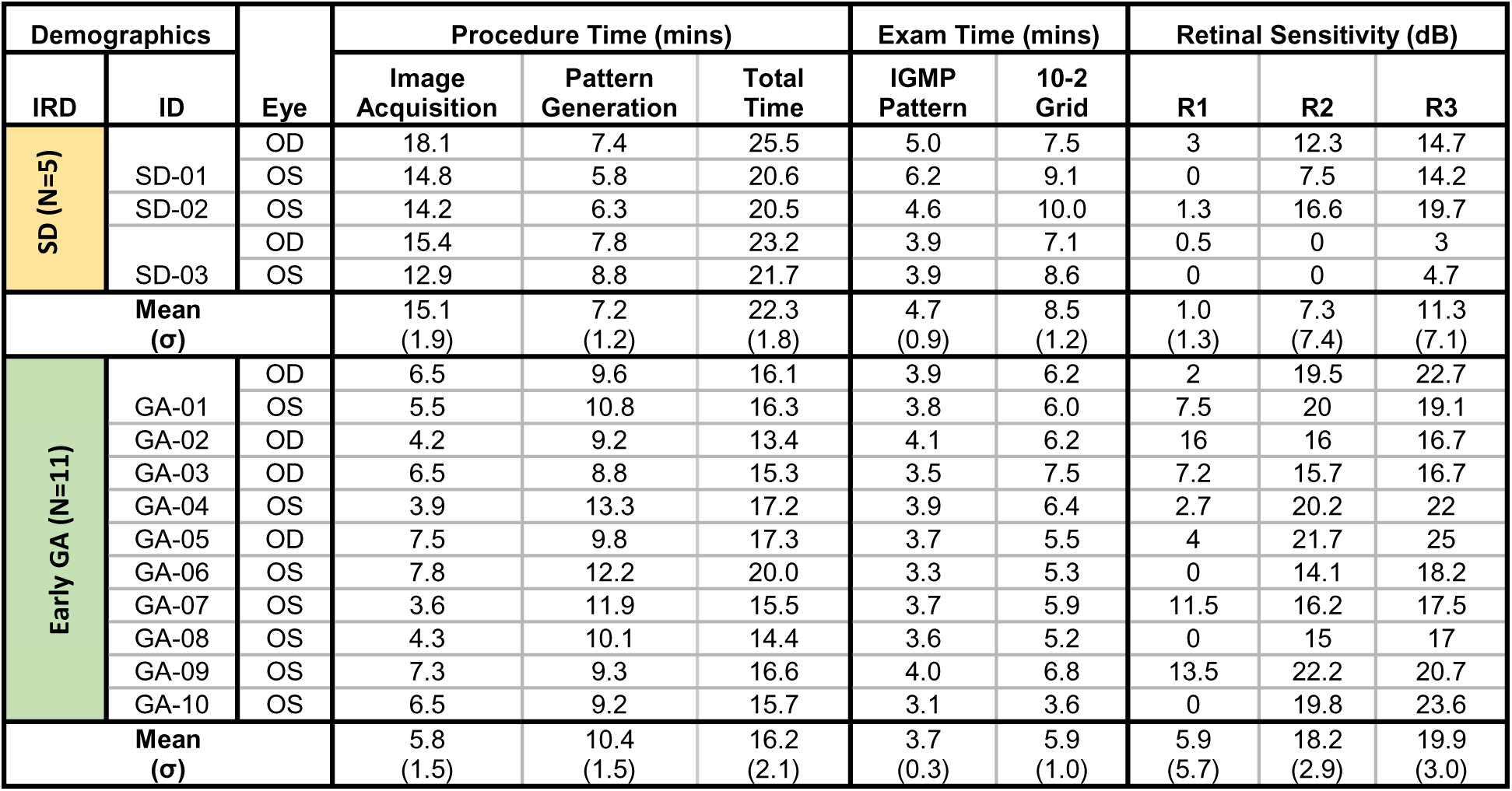
Baseline demographics, exam times, and retinal sensitivity of patients with early Geographic Atrophy and Stargardt Disease exclusively tested on the MAIA device for this study.

On average, the targeted IGMP pattern (4.7±0.9 mins) took almost half less time than standard 10-2 grid (8.5±1.2 mins) for lesion mapping in SD functional assessment. In fact, the longest duration for a targeted IGMP pattern exam to assess SD lesions was 6.2 mins which was still less than the shortest time taken for a standard 10-2 grid exam which was 7.1 mins. Similar findings were reported for eyes with GA assessed on MAIA, whereby the custom IGMP pattern took less time on average than the standard 10-2 grid. Additionally, GA lesions generally showed shorter exam durations for both MP approaches than SD lesions. In GA, the mean exam time for a targeted versus standard microperimetry pattern on MAIA was 3.7±0.3 mins versus 5.9±1.0 mins, respectively. The longest duration for a targeted IGMP pattern exam was 4.1 mins, which was comparable to the shortest time taken for a standard 10-2 grid exam, i.e. 3.6 minutes.

Additionally, in SD lesions, the mean retinal sensitivity was 1.0±1.3 dB within DDAF, 7.3±7.4 dB in TZ in QDAF, and 11.3±7.1 dB in healthy retina. The mean retinal sensitivity was higher for GA lesions than SD lesions tested on MAIA, as it was 5.9±5.7 dB in the atrophic region, 18.2±2.9 dB in the TZ, and 19.9±3.0 dB in the healthy retina. Intuitively, the mean retinal sensitivity showed an improving trend with increased distance from the degenerative lesion for all eyes, except for two cases in GA and two in SD. Of these anomalies, SD-02 showed a decrease in retinal sensitivity from R1 to R2, GA-01/OS and GA-09/OS showed a decrease in retinal sensitivity from R2 to R3, while SD-03 and GA-02 showed no change in retinal sensitivity from R1 to R2.

**Table 2** summarizes key findings of subjects with bilateral GA enrolled for MP exam on Nidek, totaling to 8 eyes. The mean time for image acquisition was 11.1±2.9 mins while custom pattern generation took an average of 18.2±4.2 mins, accounting for a total mean procedure time of 29.3±5.1 mins for GA lesions mapped via the custom IGMP approach on Nidek. This implies that the total procedure time, including both of its constituent phases, i.e. image acquisition and pattern generation, took longer on Nidek than on MAIA. Moreover, the IGMP pattern took 3.6±1.2 mins and was more timesaving than the standard 10-2 approach which took 5.8±2.0 mins when average values were considered. There was an improving trend in the mean retinal sensitivity of lesions with increased distance from atrophy for all GA lesions tested on Nidek, with R1 showing 2.3±2.5 dB, R2 showing 17.5±6.1 dB, and R3 showing 21.5±3.9 dB on the Nidek device. However, MAIA showed better results in terms of markedly higher retinal sensitivity for the same GA lesions (GA-09/OS and GA-10/OS) assessed on Nidek and when mean values were considered. Hence, our results imply that while Nidek may be more accurate, MAIA shows more sensitive readings.

**Table 2:** Baseline demographics, exam times, and retinal sensitivity of patients with early Geographic Atrophy exclusively tested on the Nidek device for this study.

| Demographics |  | Eye | Procedure Time (mins) |  |  | Exam Time (mins) |  | Retinal Sensitivity (dB) |  |  |
| --- | --- | --- | --- | --- | --- | --- | --- | --- | --- | --- |
| IRD | ID |  | Image Acquisition | Pattern Generation | Total Time | IGMP Pattern | 10-2 Grid | R1 | R2 | R3 |
| Early GA (N=8) | GA-09 | OD | 8.7 | 14.3 | 23 | 3.4 | 5.6 | 2.8 | 22.3 | 24.5 |
|  |  | OS | 10.2 | 17.2 | 27.4 | 5.6 | 8.9 | 3.5 | 20.2 | 20.8 |
|  | GA-10 | OD | 9.1 | 14.9 | 24.0 | 4.9 | 8.1 | 0 | 4.3 | 13.6 |
|  |  | OS | 8.4 | 14.5 | 22.9 | 2.3 | 3.5 | 3.2 | 22.4 | 24.8 |
|  | GA-11 | OD | 14.5 | 23.2 | 37.7 | 2.8 | 4.9 | 7.2 | 18.7 | 22.3 |
|  |  | OS | 16.3 | 25.4 | 41.7 | 2.3 | 3.8 | 0 | 14.8 | 19.2 |
|  | GA-12 | OD | 11.4 | 19.6 | 31.0 | 4.6 | 7.2 | 1.8 | 15.8 | 21.7 |
|  |  | OS | 9.8 | 16.1 | 25.9 | 3.1 | 4.6 | 0 | 21.7 | 25.4 |
| Mean (σ) |  |  | 11.1 (2.9) | 18.2 (4.2) | 29.3 (5.1) | 3.6 (1.2) | 5.8 (2.0) | 2.3 (2.5) | 17.5 (6.1) | 21.5 (3.9) |

**Table 3** summarizes key findings from the comparison of MAIA versus Nidek regarding procedure time, exam time, approach feasibility, and algorithm compatibility of custom IGMP tested on eyes with GA. The targeted pattern procedure prior to IGMP was lengthier for Nidek (29.2±7.1 mins) than MAIA (17.4±4.3 mins) although most IGMP assessments took longer on MAIA (4.0±1.0 mins) than on Nidek (3.6±1.2 mins) when mean values were considered. As reported by imaging specialists, the custom IGMP approach is feasible (100.0%) and the algorithm is compatible (100.0%) on both MP devices for all subjects.

**Table 3:** Comparing inter-operator procedure and exam times, algorithm compatibility, and approach feasibility of custom image-guided microperimetry on MAIA and Nidek devices in patients with early Geographic Atrophy.

| IRD | ID | Eye | Procedure Time (mins) |  | Exam Time (mins) |  | Is the Approach Feasible? |  | Is the Algorithm Compatible? |  |
| --- | --- | --- | --- | --- | --- | --- | --- | --- | --- | --- |
|  |  |  | Nidek | MAIA | Nidek | MAIA | Nidek | MAIA | Nidek | MAIA |
| Early GA (N=8) | GA-09 | OD | 23 | 12.5 | 3.4 | 3.7 | Y | Y | Y | Y |
|  |  | OS | 27.4 | 16.6 | 5.6 | 4.0 | Y | Y | Y | Y |
|  | GA-10 | OD | 24 | 13.2 | 4.9 | 5.3 | Y | Y | Y | Y |
|  |  | OS | 22.9 | 15.7 | 9.2 | 3.1 | Y | Y | Y | Y |
|  | GA-11 | OD | 37.7 | 24.3 | 2.8 | 5.5 | Y | Y | Y | Y |
|  |  | OS | 41.7 | 22.8 | 2.3 | 4.4 | Y | Y | Y | Y |
|  | GA-12 | OD | 31 | 18.7 | 4.6 | 2.9 | Y | Y | Y | Y |
|  |  | OS | 25.9 | 15.6 | 3.1 | 3.1 | Y | Y | Y | Y |
| Mean (σ)<br>or N (%) |  |  | 29.2<br>(7.1) | 17.4<br>(4.3) | 3.6<br>(1.2) | 4.0<br>(1.0) | Yes<br>8 (100.0) | Yes<br>8 (100.0) | Yes<br>8 (100.0) | Yes<br>8 (100.0) |

## Discussion

The transition zone (TZ) in retinal degeneration is a focal point of research due to its high disease activity and therapeutic potential. Although, this region is best visualized using FAF and OCT^20^, the slow progression of atrophic lesions^10,21^ and the lack of precise functional data limit its utility for early disease monitoring and treatment response assessment. In fact, tracking lesion growth alone prioritizes end-stage intervention over early therapeutic strategies. Moreover, given the variability in the definition of TZ across the ophthalmic literature, adopting a more restrictive definition than 500 microns (e.g., 300 microns or a pathology-based definition such as the Ellipsoid Zone Defect) would further limit and non-uniformly distribute the testing points when using the 10-2 pattern, thereby diminishing the significance of the findings.

As **summarized by our key findings and** illustrated in **Figure 9**, targeted IGMP thus offers several key methodological advantages over the standard 10-2 grid. The 10-2 pattern samples the TZ asymmetrically, resulting in variable—and sometimes suboptimal—coverage. In contrast, the custom IGMP pattern evenly distributes the test points while targeting TZ, thereby increasing spatial resolution and consistency of sampling at the lesion margins. Targeted IGMP thus reduces sampling bias, potentially yielding more reliable data for monitoring lesion progression over time. The custom approach can potentially enhance sensitivity to subtle functional changes at the lesion margin and improve reproducibility in longitudinal assessments by ensuring uniform testing of the TZ across subjects and cohorts. When combined with the method’s adaptability to varying lesion sizes and its potential for tailoring the TZ definition, this approach establishes a more robust framework for evaluating GA and related macular pathologies in clinical research settings.

**Figure 9:**
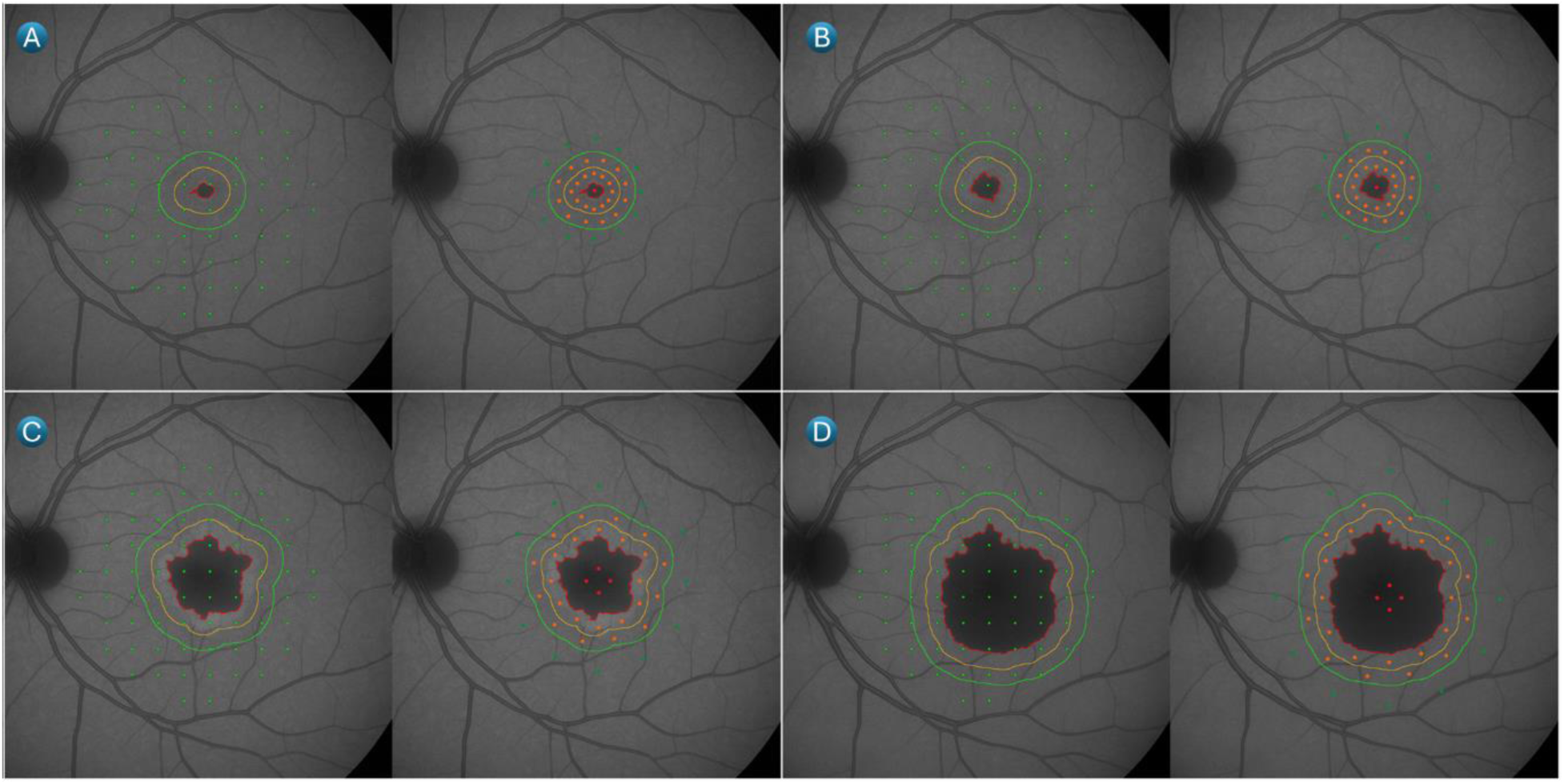
Representative artificially generated Fundus Autofluorescence images illustrating areas of definitely decreased autofluorescence (DDAF) analogous to varying sizes of geographic atrophy. For each lesion **(A–D)**, the left image shows a superimposed standard 10-2 microperimetry test point distribution (green dots), while the right image demonstrates a custom image-guide microperimetry (IGMP) pattern (orange dots) specifically targeting the transitional zones (TZ) of the lesion, defined as 500 microns away from DDAF. **(C)** shows that the standard pattern provides only seven test points within TZ, many of which are positioned too close to the lesion or the outer boundary, whereas the custom pattern ensures the placement of 12 uniformly distributed orange test points between the red (inner) and yellow (outer) outlines, offering a more representative assessment of TZ health. This disparity is even more pronounced in smaller lesions, as seen in **(A)**, where the 10-2 pattern places just two points (with only one optimally positioned), compared to 12 systematically arranged points using the targeted approach.

Using microperimetry assessments, the ProgStar studies on SD found that a 1-dB decline in MP readings correlated to a 0.59 ETDRS letter loss for BCVA monitoring^19,22,23^. Similarly, a phase 1/2 trial on choroideremia also detected a greater decline in mean retinal sensitivity in untreated vs. treated eyes using microperimetry^24–26^. Our proof-of concept with targeted image-guided microperimetry thus holds transformative potential for advancing our understanding and treatment of retinal diseases like GA^9,13^, SD^22,23^, and diabetic macular ischemia (DMI)^33,34^. By enabling precise correlation between structural changes in the retina and their functional impacts, IGMP can significantly improve the precision needed to detect subtle, early changes in retinal function that may not be visible through traditional imaging techniques^12,35,36^. This capability is crucial for identifying potential intervention points before significant damage occurs, opening new avenues for treatments aimed at neuroprotection.

Recently, the Food and Drug Administration (FDA) approved pegcetacoplan, a complement C3 inhibitor, which reduced GA lesion growth by 36% with monthly injections in the OAKS & DERBY trials^3^, though its impact on functional vision remains uncertain. Therefore, multiple studies are now investigating microperimetry as a functional endpoint for therapeutic targets of IRDs^17,22,27–30^. Our findings establish the feasibility of implementing a targeted IGMP approach in atrophic retinal diseases such as GA, where TZ can be precisely delineated from OCT imaging, and SD, where the atrophy can be segmented from FAF imaging. This approach allows for a more focused functional assessment in clinically relevant regions. Additionally, our study highlights the potential for improved testing efficiency, as evidenced by the significant reduction in examination time, which may enhance the practicality and integration of microperimetry into clinical and research settings. This aligns with the PINNACLE Study Report-7^37^, which underscores the limitations of standard microperimetry grids in detecting localized retinal sensitivity, especially in lesions with ellipsoid and/or interdigitation loss and subretinal fluid retention, leading to overestimations in functional assessment for eyes with intermediate age-related macular degermation (iAMD).

Our approach thus aimed to address these challenges by ensuring that microperimetry targets clinically relevant regions identified through OCT and FAF imaging, thereby improving the precision of structure-function correlation. We demonstrate the feasibility of an image-integrated targeted microperimetry workflow, successfully implementing custom software to significantly reduce examination time while maintaining precise functional assessments. The substantial reduction in testing time may increase patient compliance and the practicality of microperimetry in clinical and research settings. Limitations include a small sample size, particularly with Nidek- MP3/S, and initial testing limited to mainly GA and SD. Validation in larger, more diverse cohorts and application to other retinal diseases is required.

Future studies employing the custom image-based microperimetry on larger scale will allow for more precise visual function assessment and increase the yield of microperimetry for regulatory requirements while keeping the duration of the study at a minimum by reducing the time and resources required to conduct microperimetry. Future studies should also aim to evaluate method reliability and repeatability, compare targeted microperimetry sensitivity against standard protocols, and investigate correlations with other biomarkers. Refinement of the automated pattern-generation process could further enhance accessibility. In conclusion, this targeted microperimetry workflow presents a feasible and efficient method for detailed functional assessment in retinal diseases, with potential to significantly improve disease monitoring and therapeutic evaluation.

## Data Availability

All data produced in the present study are available upon reasonable request to the authors.

## Acknowledgments

The authors would like to acknowledge research volunteers, mainly Aly Hamza Khowaja, at the Sepah Lab in the Byers Eye Institute for their contribution in data collection and analysis.

## Author Contributions

YJS, RA, and MSH conceptualized the study. HY and MC executed and streamlined the study methods. RA, ZZF, and KJ collected the data. HY and MC coded the MATLAB application. MA, ZZF and YJS analyzed and presented the data. MSH, QDN and SMS composed the initial manuscript. All authors reviewed and approved the final draft.

## Meeting Presentation

The abstract for this manuscript has not been presented at any conference.

## Author Contacts

Rubbia Afridi (650) 723-6995, M Sohail Halim (650) 723-6995, Mohamed Ahmed (669) 699-1080, Hikmet Yucel (669) 699-1080, Zoha Zahid Fazal (650) 723-6995, Kholood Janjua (650) 723-6995, Mauro Campigotto (669) 699-1080, Syed Mahmood (608) 782-7300, Quan Dong Nguyen (650) 723-6995, Yasir Jamal Sepah (650) 723-6995

## Abbreviations List

BCVA: Best-Corrected Visual Acuity
CFP: Color Fundus Photo
cRORA: complete Retinal Pigment Epithelium Outer Retinal Atrophy
DDAF: Definitely Decreased Autofluorescence
DMI: Diabetic Macular Ischemia
FAF: Fundus Autofluorescence
FDA: Food and Drug Administration
GA: Geographic Atrophy
GUI: Graphical User Interface
HS: Heidelberg Spectralis
iAMD: intermediate age-related macular degeneration
IGMP: Image-Guided Microperimetry
IR: Infrared
IRD: Inherited Retinal Disease
MAIA: Macular Integrity Assessment system
MATLAB: Matrix Laboratory
MCS: Medium Coordinate System
MP: Microperimetry
MP3/S: Microperimeter MP3 Type S by Nidek
NEI: National Eye Institute
OCT: Optical Coherence Tomography
OD: Right eye
OS: Left eye
PRL: Preferred Retinal Locus
PCS: Perimetric Coordinate System
QDAF: Questionably Decreased Autofluorescence
R1: Ring 1 for IGMP grid; bounds the atrophic lesion inside
R2: Ring 2 for IGMP grid; bounds the transition zone inside
R3: Ring 3 for IGMP grid; bounds the healthy retina both sides
ROI: Region of Interest
RPE: Retinal Pigment Epithelium
S1: Scan 1 for pre-MP protocol; infrared OCT image
S2: Scan 2 for pre-MP protocol; enface OCT + B-scan
S3: Scan 3 for pre-MP protocol; annotated OCT image
SN1: Scan N1; raw IR enface image from MAIA
SM1: Scan M1; CFP raw export from Nidek-MP3/S
SD: Stargardt Disease
SURF: Speeded-Up Robust Features
TZ: Transition Zone
XML: Extensible Markup Language

